# Machine Learning-Based Hyperglycemia Prediction: Enhancing Risk Assessment in a Cohort of Undiagnosed Individuals

**DOI:** 10.1101/2023.11.22.23298939

**Authors:** Kolapo Oyebola, Funmilayo Ligali, Afolabi Owoloye, Blessing Erinwusi, Yetunde Alo, Adesola Musa, Oluwagbemiga Aina, Babatunde Salako

**Affiliations:** Centre for Genomic Research in Biomedicine, Mountain Top University, Ibafo, Nigeria; Nigerian Institute of Medical Research, Lagos, Nigeria; Habilis Biotech Limited, Nigeria

**Keywords:** Hyperglycemia, Diabetes, Machine Learning, Hypertension, Random Forest

## Abstract

**Background:** Noncommunicable diseases (NCDs) continue to pose a significant health challenge globally, with hyperglycemia serving as a prominent indicator of potential diabetes. This study employed machine learning algorithms to predict hyperglycemia in a cohort of asymptomatic individuals and unraveled crucial predictors contributing to early risk identification.

**Methods:** This dataset included an extensive array of clinical and demographic data obtained from 195 asymptomatic adults residing in a suburban community in Nigeria. The study conducted a thorough comparison of multiple machine learning algorithms to ascertain the most effective model for predicting hyperglycemia. Moreover, we explored feature importance to pinpoint correlates of high blood glucose levels within the cohort.

**Results:** Elevated blood pressure and prehypertension were recorded in 8 (4%) and 18 (9%) individuals respectively. Forty-one (21%) individuals presented with hypertension (HTN), of which 34/41 (82.9%) were females. However, cohort-based gender adjustment showed that 34/118 (28.81%) females and 7/77 (9.02%) males were hypertensive. Age-based analysis revealed an inverse relationship between normotension and age (r = -0.88; P < 0.05). Conversely HTN increased with age (r = 0.53; P < 0.05), peaking between 50-59 years. Isolated systolic hypertension (ISH) and isolated diastolic hypertension (IDH) were recorded in 16/195 (8.21%) and 15/195 (7.69%) individuals respectively, with females recording higher prevalence of ISH 11/16 (68.75%) while males reported a higher prevalence of IDH 11/15 (73.33%). Following class rebalancing, random forest classifier gave the best performance (Accuracy Score = 0.894; receiver operating characteristic-area under the curve (ROC-AUC) score = 0.893; F1 Score = 0.894) of the 27 model classifiers. The feature selection model identified uric acid and age as pivotal variables associated with hyperglycemia.

**Conclusions:** Random Forest classifier identified significant clinical correlates associated with hyperglycemia, offering valuable insights for early detection of diabetes and informing the design and deployment of therapeutic interventions. However, to achieve a more comprehensive understanding of each feature’s contribution to blood glucose levels, modeling additional relevant clinical features in larger datasets could be beneficial.

## Introduction

Non-communicable diseases (NCDs) have become a significant public health concern in Africa [1]. Conditions like coronary artery disease, stroke, hypertension, and diabetes, which were once primarily associated with developed nations or affluence, have now become pervasive health challenges in developing countries and across diverse socio-economic strata [1]. The complex nature of NCDs underscores the need for a comprehensive approach to risk assessment, intervention and prevention.

Suburban communities serve as a distinctive microcosm within an evolving landscape of diseases [2, 3]. These communities, characterized by the coexistence of traditional and modern lifestyles, grapple with risk factors that necessitate thorough examination [4]. The epidemiological shift from communicable to non-communicable diseases, coupled with limited healthcare resources especially in suburban parts of developing countries [5, 6], stresses the importance of this research. In addition, recent advancements in genetic research have elucidated the underlying mechanisms of various complex NCDs. The identification of individuals at an elevated genetic risk for NCDs has the potential to revolutionize the approach of healthcare stakeholders to disease management. However, the effective implementation of genetic screening for NCD risk analysis relies on a robust understanding of the baseline contributors prevalent in the target population [7, 8]. This study provided a comprehensive description of the prevalence and intricate interplay of risk factors associated with NCDs, highlighting hypertension, obesity and diabetes. The specific focus was on undiagnosed asymptomatic individuals to elucidate the complex relationships of these health indicators within this population.

Machine learning (ML) encompasses a diverse set of algorithms designed to extract patterns from data and establish associations between these patterns and discrete sample classes within the data. ML proves to be a valuable tool for identifying potential disease risk factors, elucidating etiology and interpreting complex pathological processes in the context of NCDs [9–11]. In this study, multiple ML algorithms were developed to predict elevated blood glucose levels in a cohort of undiagnosed asymptomatic individuals. The primary objective was to systematically compare the accuracies of supervised machine learning classifiers to identify the most effective model for predicting hyperglycemia. Leveraging the predictors in the dataset, we meticulously constructed and evaluated these models for the identification of significant features associated with potential diabetes in the population.

This research serves as a critical foundation in advancing our understanding of the evolving landscape of NCDs. Furthermore, it contributes to the progression of precision medicine approaches in disease management by leveraging the capabilities of machine learning in unraveling the intricate dynamics of hyperglycemia prediction and potentially informing tailored interventions for individuals at risk.

## Methods

### Participant recruitment and screening

This study was carried out as part of a parallel community-based genetic screening of apparently healthy adults living in Ijede Community, Lagos, Nigeria. Ethical approval was obtained from the Institutional Review Board of the Nigerian Institute of Medical Research (IRB/21/074). Following informed consent, participants were recruited and 10ml of venous blood samples were collected per individual. Demographic information, body mass index (BMI), knowledge, attitude and practices were obtained from the participants. The study clinician further clerked participants for personal and family medical history as well as their smoking status. Exclusion criteria included pregnancy at the time of recruitment, placement on antihypertensive or antidiabetic chemotherapy, radiotherapy, current or previous hematologic or tumoral diseases and known chronic diseases. Participants underwent electrocardiogram (ECG) screening (SonoHealth, USA) to provide clues on heart defects or other heart-related problems. Hemoglobin electrophoresis was conducted to detect possible hemoglobinopathy in the participants [12]. In addition, random blood glucose concentrations (Guilin Royalze, China) and blood pressure (BP) values (Iston Mediq, USA) were determined to evaluate the presence or absence of prediabetes, diabetes, prehypertension (preHTN) or hypertension (HTN) onset in the participants. Individuals with screening tests outside normal ranges were advised to visit their healthcare specialists for further checks. Normal BP was described as systolic blood pressure (SBP) <120mmHg and diastolic blood pressure (DBP) <80 mmHg. Elevated BP was defined as SBP of 120–129 mmHg and DBP <80 mmHg, stage 1 hypertension (preHTN) as SBP ≥ 130-139 mmHg and DBP 80 – 89 mmHg and stage 2 HTN as SBP ≥140 and DBP ≥ 90 mmHg [13]. Isolated systolic hypertension (ISH) was described as SBP above 140 mmHg with diastolic blood pressure (DBP) of less than 90 mmHg [14]. Isolated diastolic hypertension (IDH) is an important subtype of hypertension defined as a systolic blood pressure (SBP) of <130 mm Hg and a diastolic blood pressure (DBP) of at least 80 mm Hg [15]. Prediabetes was defined as random blood glucose (RBG) concentration of 140–199 mg/dl or fasting blood glucose of 100–125 mg/dl. Diabetes mellitus was defined as random blood glucose level of ≥200 mg/dl or fasting blood glucose of ≥126 mg/dl [16]. However, as all the participants reported they were not fasting, random blood glucose values were documented.

### Data analysis

Data cleaning, exploratory analysis and feature engineering were performed in Google Colab (with Python 3.10). The target variable was specified as “blood glucose,” where 1 indicated a RBG concentration ≥140mg/dl and 0 indicated RBG concentration <140mg/dl. Independent variables included age (integer), sex (integer), BMI (float), smoking status (integer), ECG (float), hemoglobin (float), cholesterol (float), uric acid (float), systolic blood pressure (integer), diastolic blood pressure (integer), normal BP (integer), elevated BP (integer), preHTN (integer), HTN (integer), isolated systolic hypertension (integer), isolated diastolic hypertension (integer), prediabetes (integer), diabetes (integer), normal glucose (integer), abnormal ECG values (integer) and normal ECG values (integer). The dataset was checked and visualized for missingness using seaborn heatmap (Supplementary Figure 1). Missing values were replaced with column mean (for continuous variables) or mode (for categorical variables). Duplicate rows and outliers were dropped before encoding categorical variables and creating dummy variables. Subsequently, we created a heatmap of correlation of independent variables with target column in descending order. The cleaned dataset was then scaled for subsequent training of machine learning models. P-value < 0.05 was considered statistically significant.

### Machine learning models and deployment as public API

The study adopted 27 supervised classification algorithms and compared their accuracies to identify the best performing model for predicting high blood glucose which was defined in this study as random blood glucose (RBG) concentration ≥140mg/dl (Figure 1). Specifically, after installation and importation of Sci-Kit Learn libraries [17], we carried out data cleaning, exploration and scaling to improve the efficiency of our model (Supplementary Methods). Imbalances in the distribution of hyperglycemia cases and non-cases within the dataset might affect the model’s performance. Addressing this imbalance and validating the model on balanced datasets could enhance its robustness. To address class imbalance in the outcome variable (blood glucose level), we adopted synthetic minority over-sampling technique (SMOTE). SMOTE tackled the underrepresentation of the minority class) and rebalanced the class distribution for equitability [18]. After resampling, we split the data into training and test sets at ratio 80:20 respectively, using the train_test_split function in Sci-Kit Learn. We went further to rank the performances of the machine learning algorithms using LazyPredict to obtain the weighted average of the F1 and accuracy scores as well as the receiver operating characteristic-area under the curve (ROC-AUC) score. For hyperparameter optimization, we adopted GridSearchCV (https://github.com/oyebolakolapo/Machine-Learning-Prediction-of-Elevated-Blood-Glucose-in-a-Cohort-of-Apparently-Healthy-Adults). The grid search technique constructs many versions of the model with all possible combinations of hyperparameters to return the best one [19]. Subsequently, we determined feature importance to provide insight into which features are most associated with elevated blood glucose level using the best performing model. To operationalize the best performing model generated at scale, the training file was stored as a serialized pickle file. Subsequently, we used Fast application programming interface (Fast API) in Google Colab [20], to make an inference call from the model using the predict () function and generated our API. Pyngrok was used to open secure tunnels from public URL to local host.

**Figure 1:**
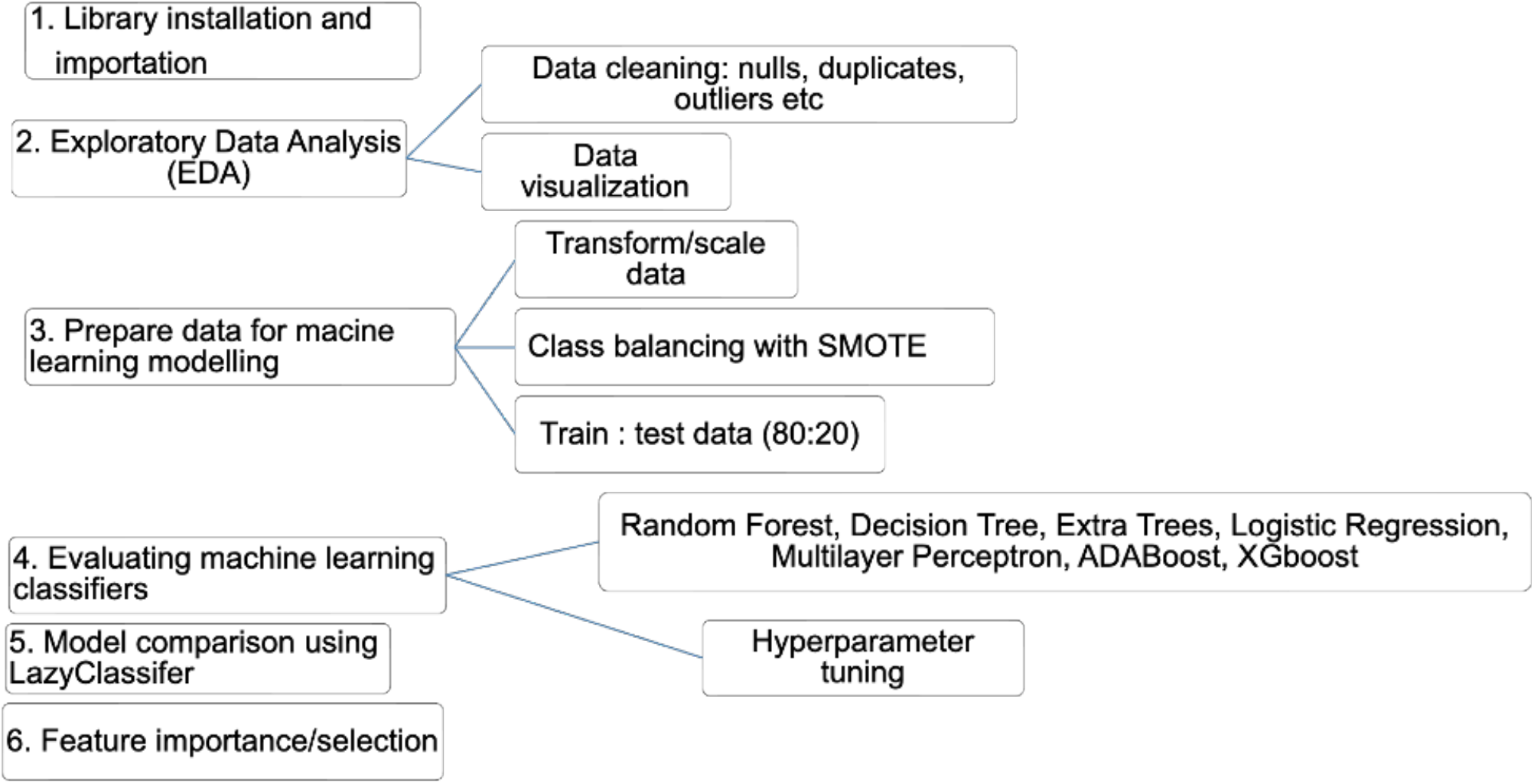
Machine learning model development.

## Results

### Cohort description

Two hundred participants aged 18-83 years were enrolled into the cohort. However, after hemoglobin electrophoresis screening, five individuals were found to possess the HbSS/HbSC genotypes and were excluded from further analysis. Enlisted individuals consisted of 118 females and 77 males (Figure 2; Supplementary Figure 2).

**Figure 2:**
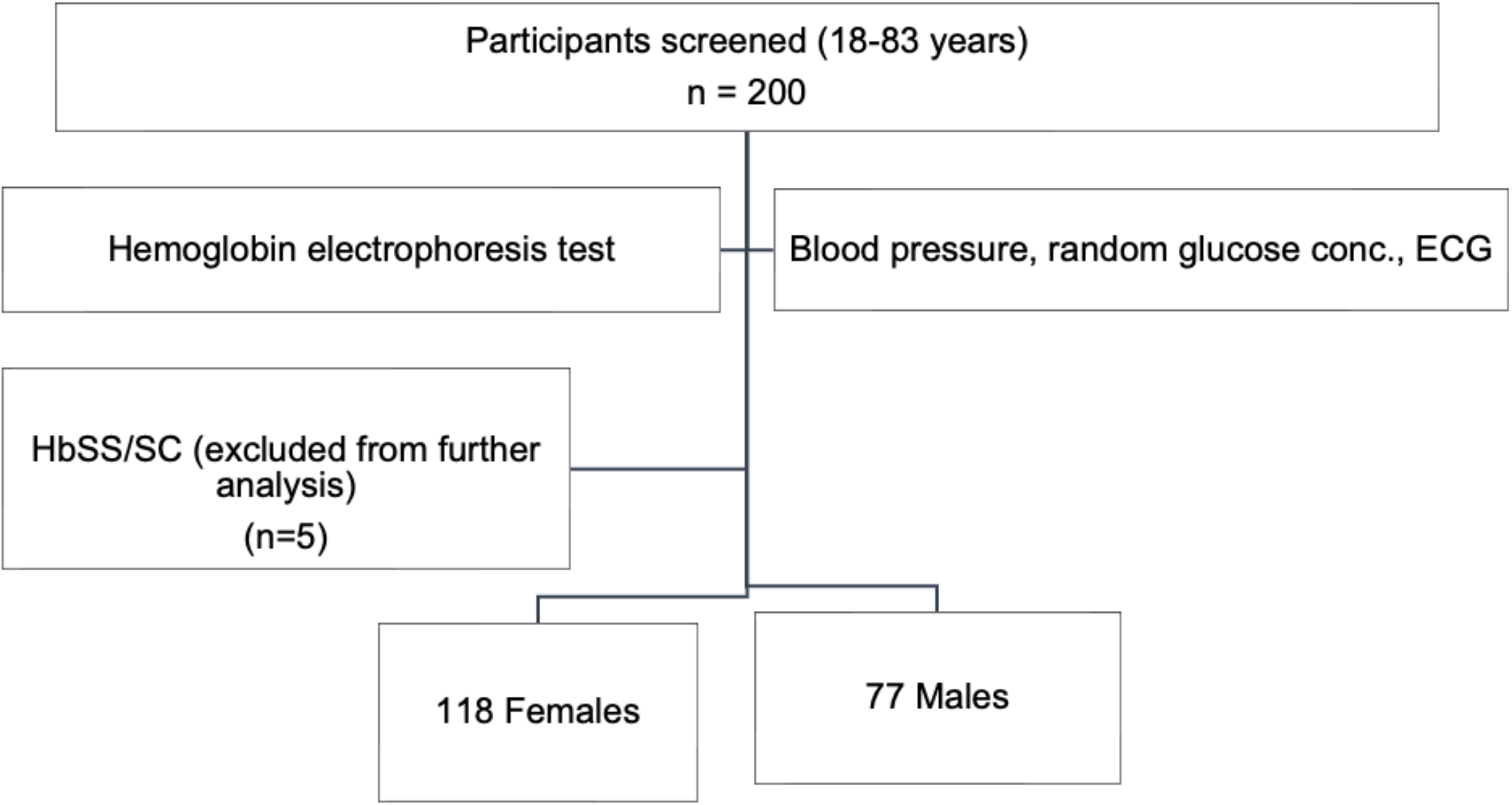
Participant recruitment and screening.

### Correlation analysis

Participants were categorized into six age groups: 18-29; 30-39; 30-49; 50-59; 60-69 and =>70 years. Elevated blood pressure and preHTN were recorded in 8 (4%) and 18 (9%) individuals respectively (Figure 3). Forty-one (21%) of the cohort (n = 195) presented with HTN, of which 34/41 (82.9%) were females (Supplementary Figure 3). Age-based analysis revealed an inverse relationship between normotension and age (r = -0.88; P < 0.05). Consistently, HTN increased with age (r = 0.53; P < 0.05), peaking between 50-59 years (Figure 4). ISH and IDH were recorded in 16/195 (8.21%) and 15/195 (7.69%) individuals respectively, with females recording higher prevalence of ISH 11/16 (68.75%) while males reported a higher prevalence of IDH 11/15 (73.33%) (Supplementary Figure 4). There was a positive correlation between ISH and participants’ age (r = 0.86; P < 0.05), whereas IDH was inversely correlated with age (r = -0.71) (Figure 5). We went further to examine the heart rates of the participants and observed an age-dependent increase in the percentage of participants with abnormal ECG values peaking between 60-69 years (Figure 6). However, no significant difference was observed in the ECG values of male and female participants (*X*^2^= 0.1257, P > 0.05) (Supplementary Figure 5). Random blood glucose value between 140 - 199mg/dl (prediabetes) was detected in 22 (11.58%) participants, while diabetes was suspected in five (2.63%) individuals (Figure 7). An inverse relationship (r = -0.81; P < 0.05) was observed between age and normal glucose level, whereas the frequency of prediabetes (r = 0.63; P < 0.05) and suspected diabetes (r = 0.58; P < 0.05) seemed to increase with age (Supplementary Figure 6). Meanwhile, a correlation matrix between each independent variable and the target column (blood glucose level) showed that age had the highest ranking even though the correlation coefficient was weak (Figure 8; Supplementary Figure 7).

**Figure 3:**
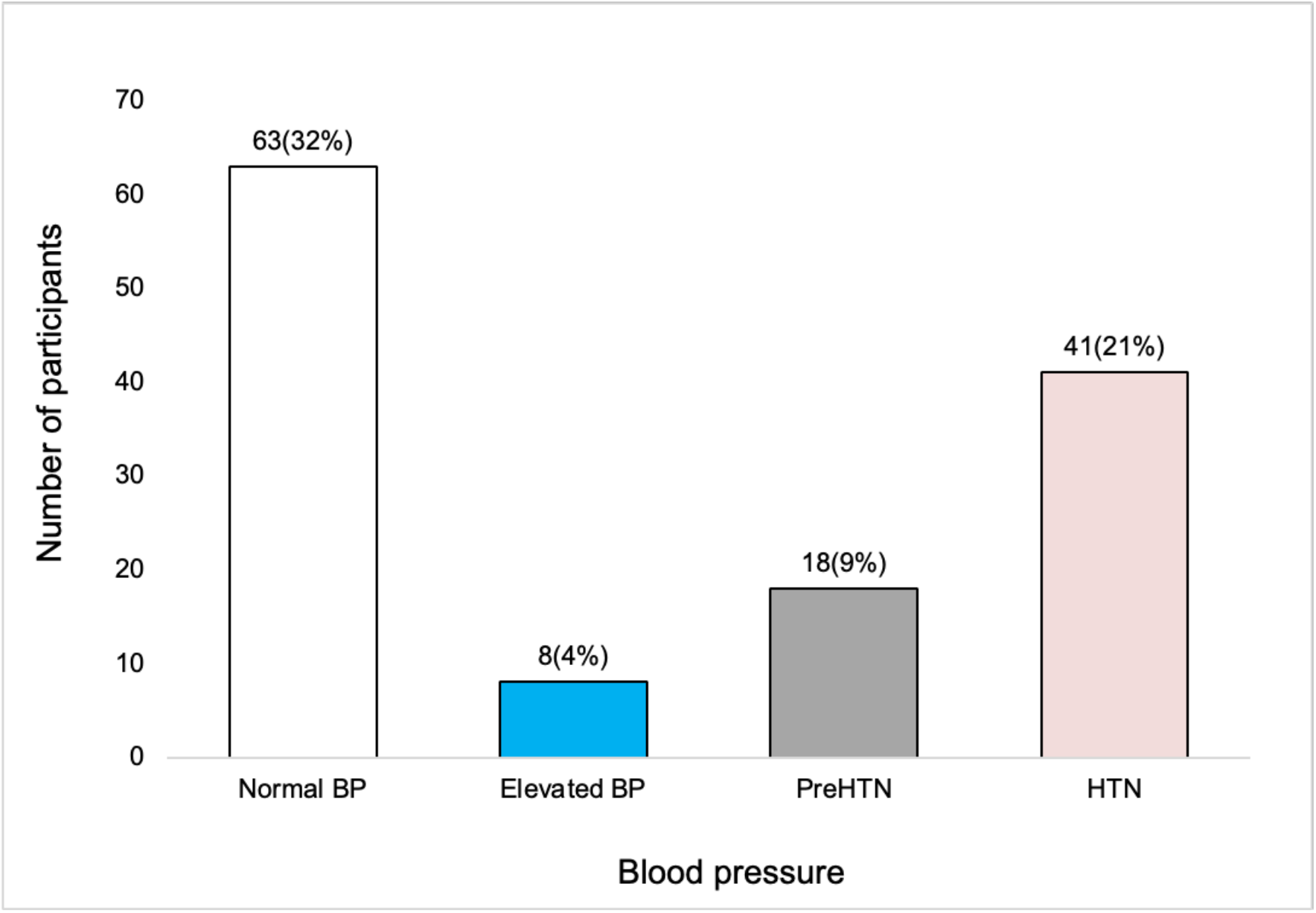
Blood pressure values in the cohort (BP = Blood Pressure; HTN = Hypertension)

**Figure 4:**
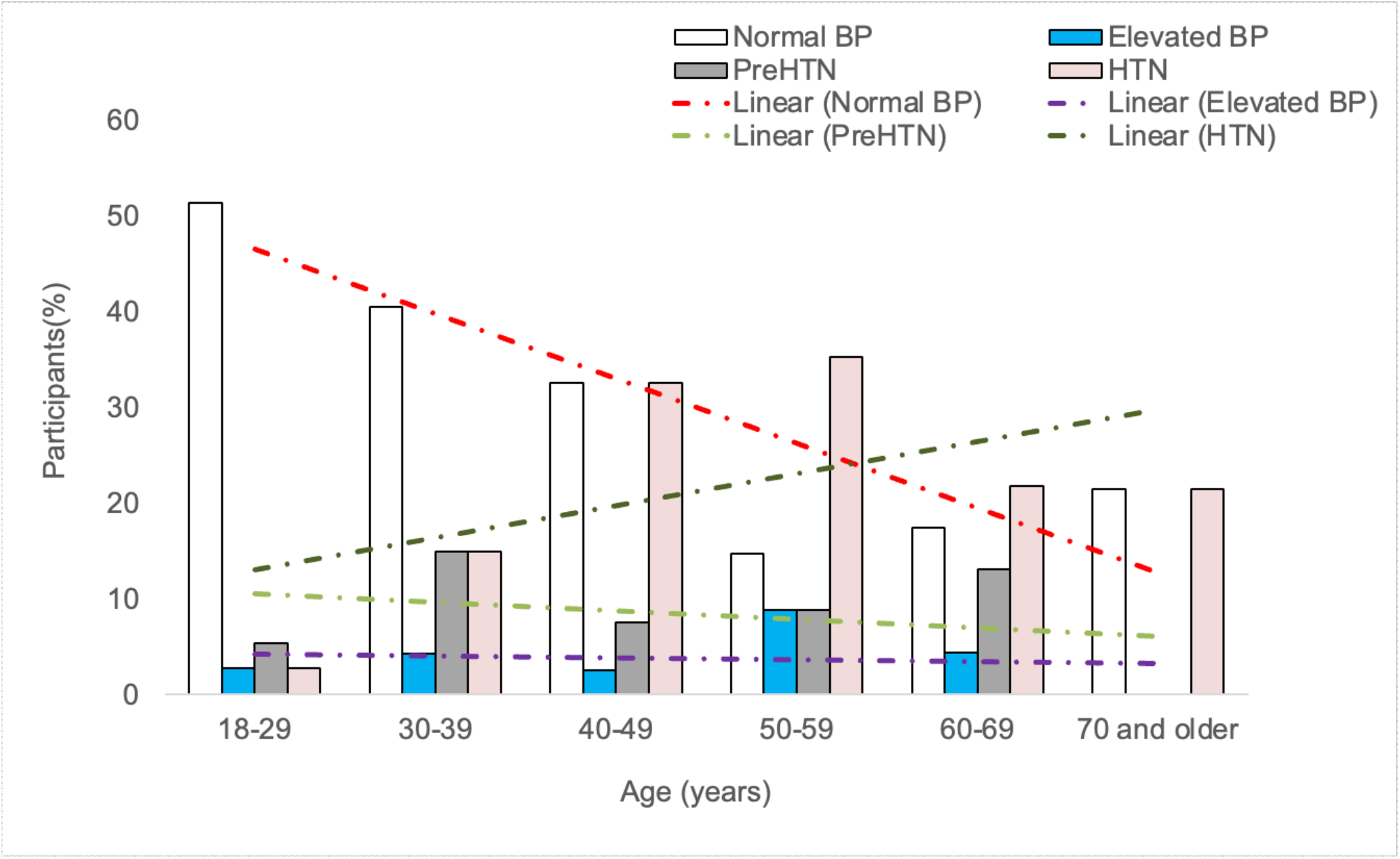
Age-based analysis of blood pressure in the cohort.

**Figure 5:**
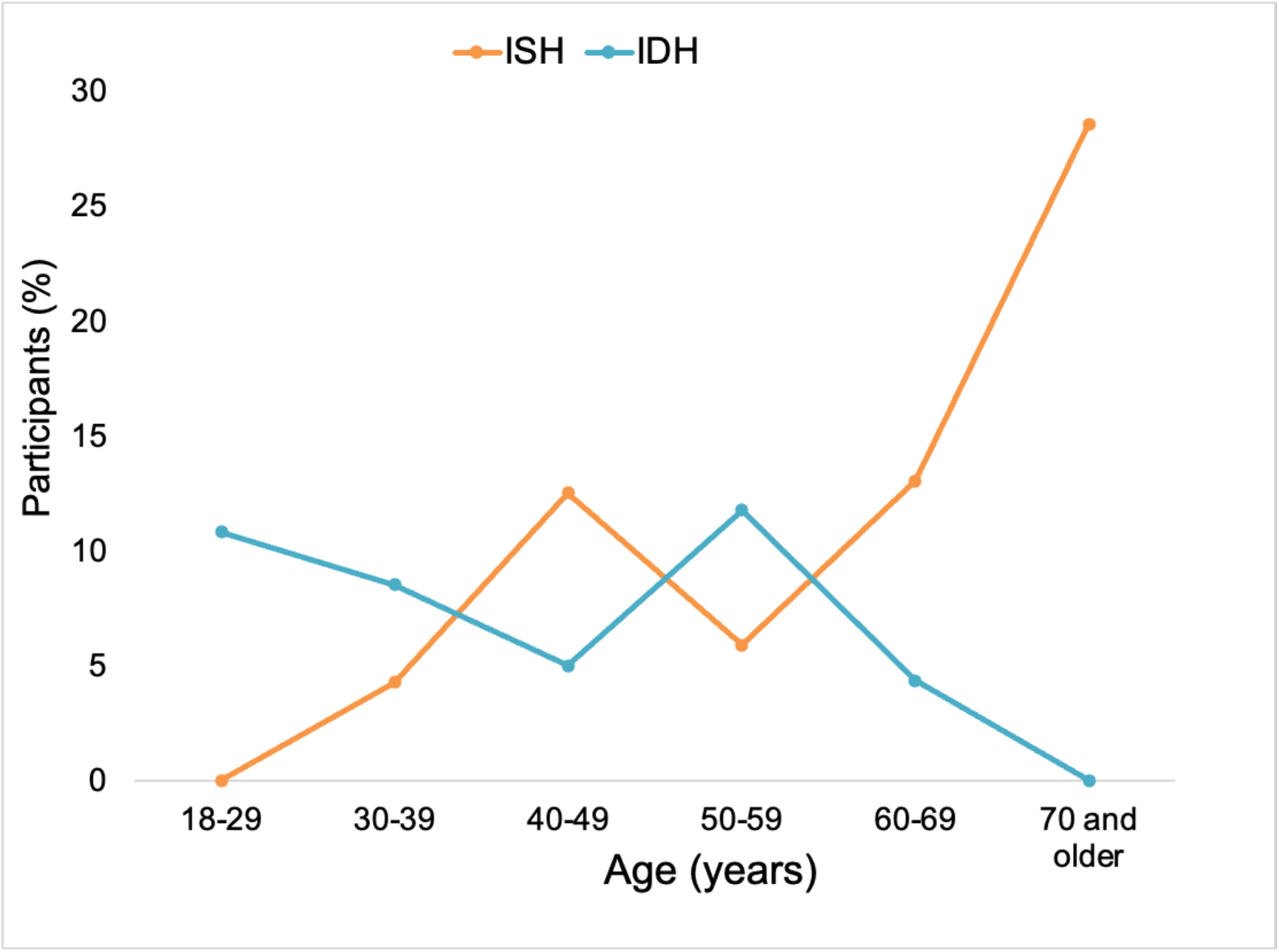
Age-based analysis of isolated systolic hypertension (ISH) and isolated diastolic hypertension (IDH) in the cohort.

**Figure 6:**
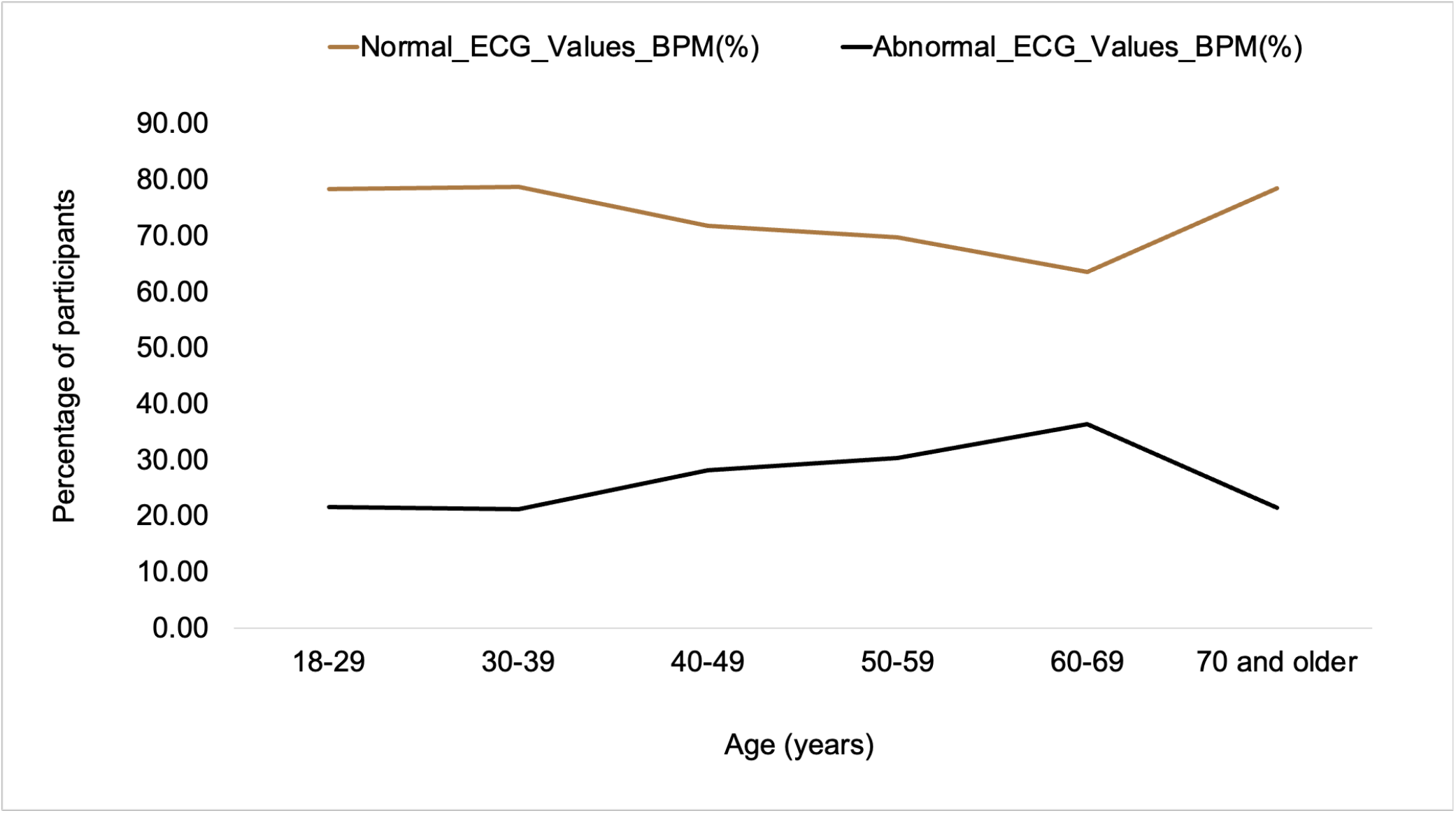
Age-based electrocardiogram (ECG) analysis.

**Figure 7:**
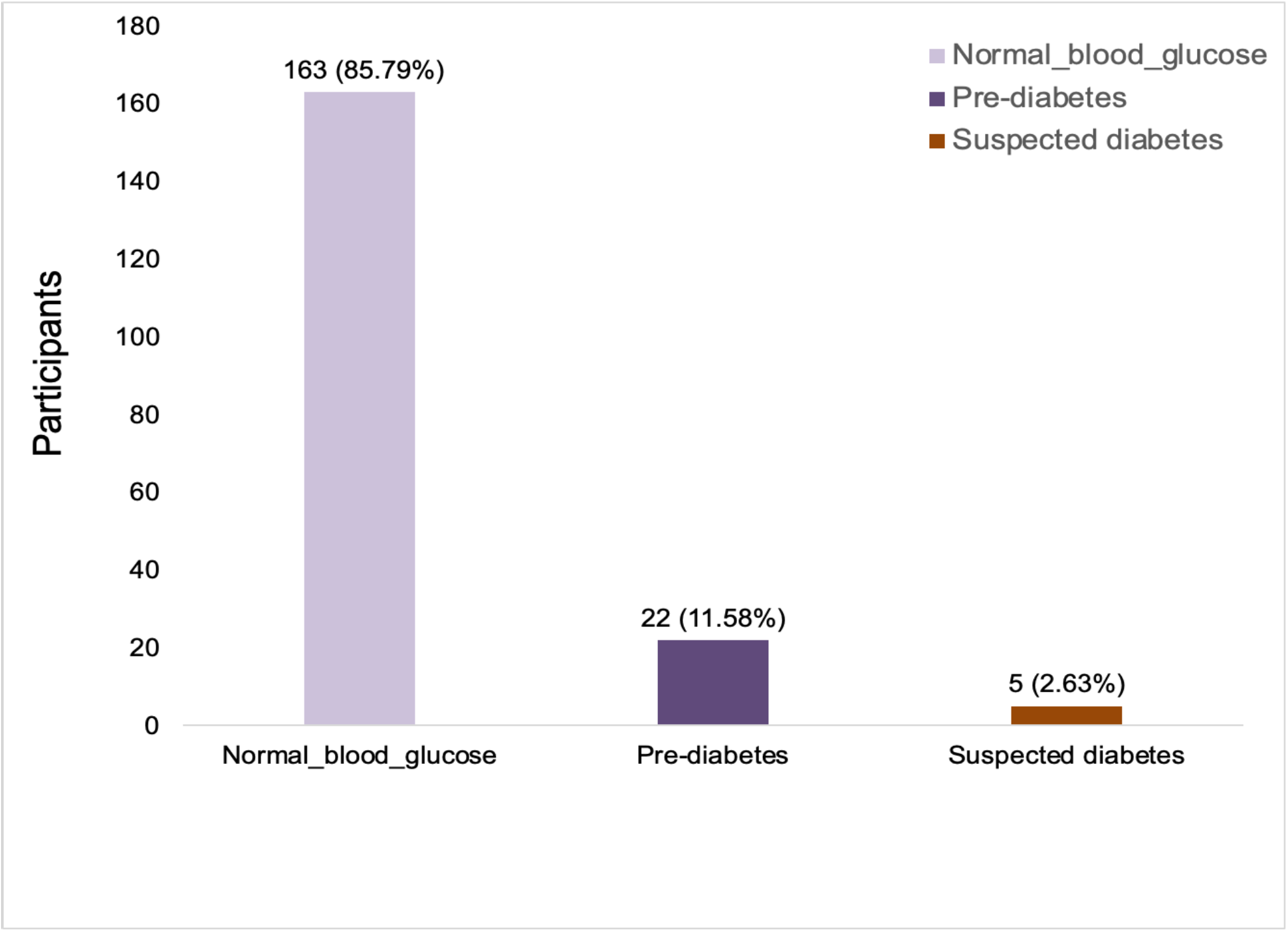
Blood glucose levels in the cohort.

**Figure 8:**
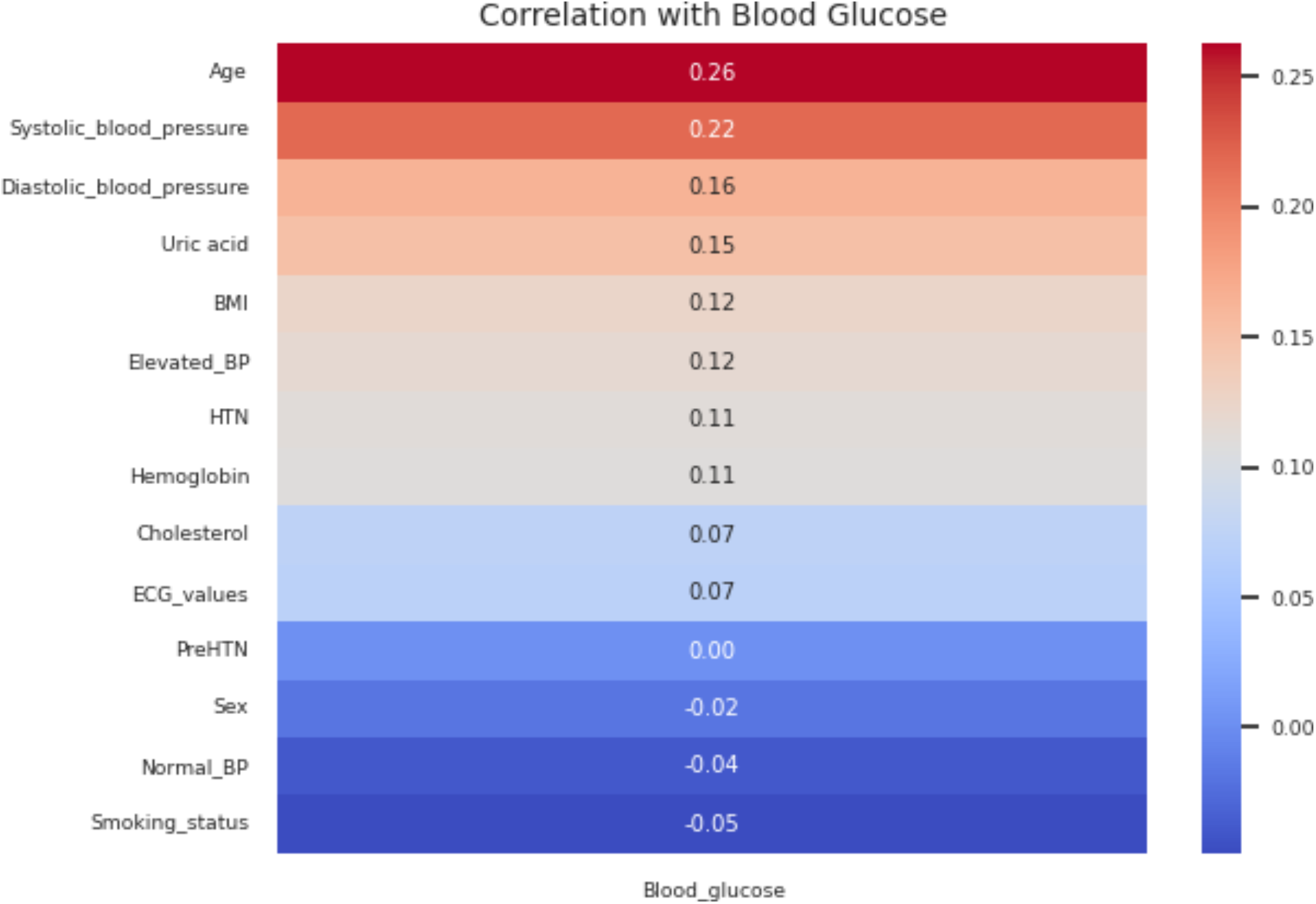
Correlation matrix of independent variables with the outcome variable.

### Model evaluation

Following data cleaning, transformation (Supplementary Figure 8) and observation of a class imbalance in the target variable (Supplementary Figure 9), whereby the raw dataset demonstrated that 83.6% of the participants had normal blood glucose {0} while 16.4% had high blood glucose level {1}, rebalancing was established with SMOTE to yield an even representation of both categories of blood glucose level (Counter ({0: 163, 1: 163}). When the performance of each classifier was tested, the reports showed Random Forest Classifier (Figures 9 and 10) gave the best accuracy (Accuracy Score = 0.894; ROC-AUC score = 0.891; F1 Score = 0.893) followed by Extra Trees (Accuracy Score = 0.879; ROC-AUC score = 0.875; F1 Score = 0.878) and XGB classifiers (Accuracy Score = 0.864; ROC-AUC score = 0.862; F1 Score = 0.863), respectively (Figure 9B; Table 1).

**Figure 9:**
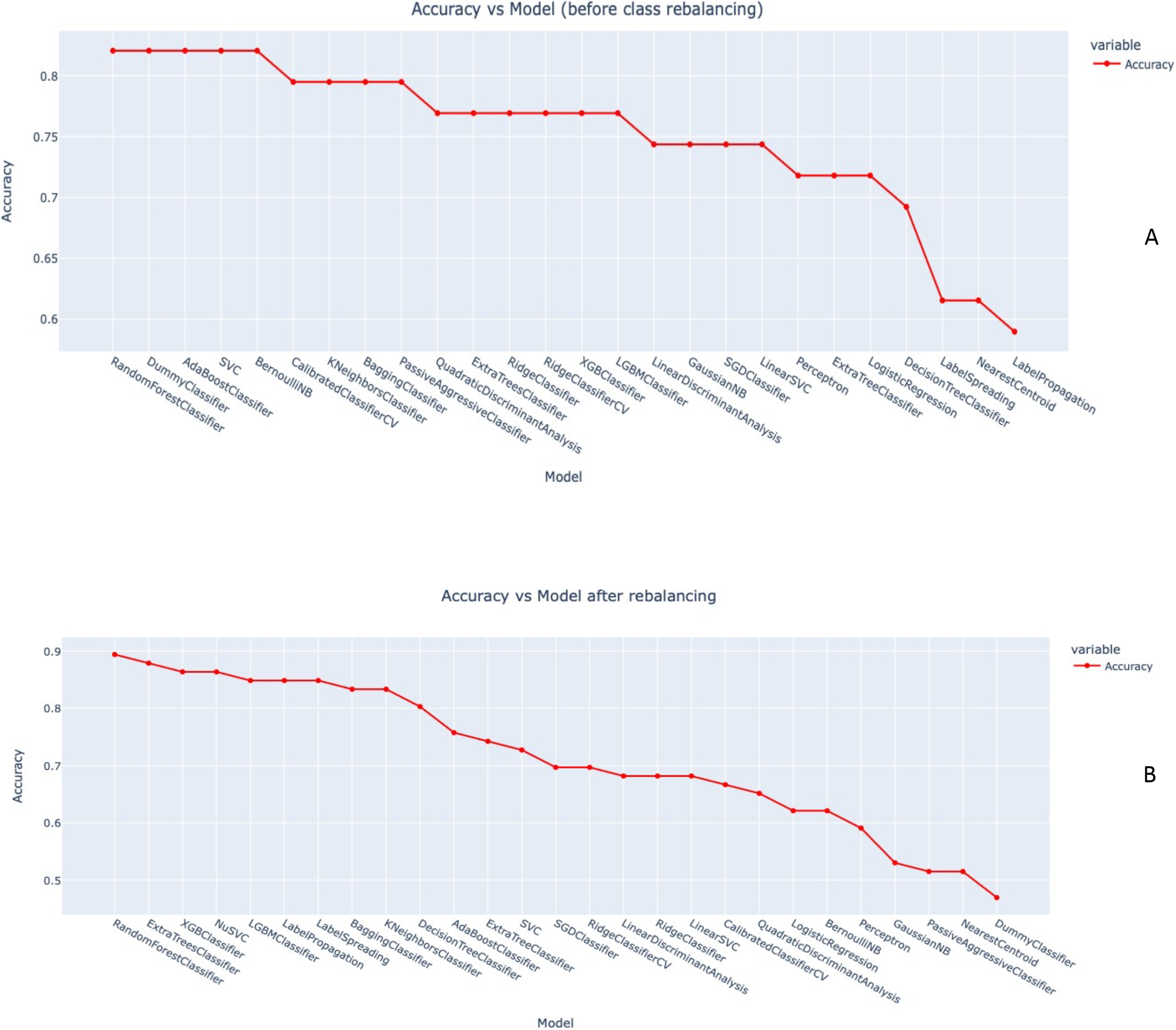
Accuracy scores of machine learning classifiers (A) before class rebalancing with SMOTE.

**Figure 10:**
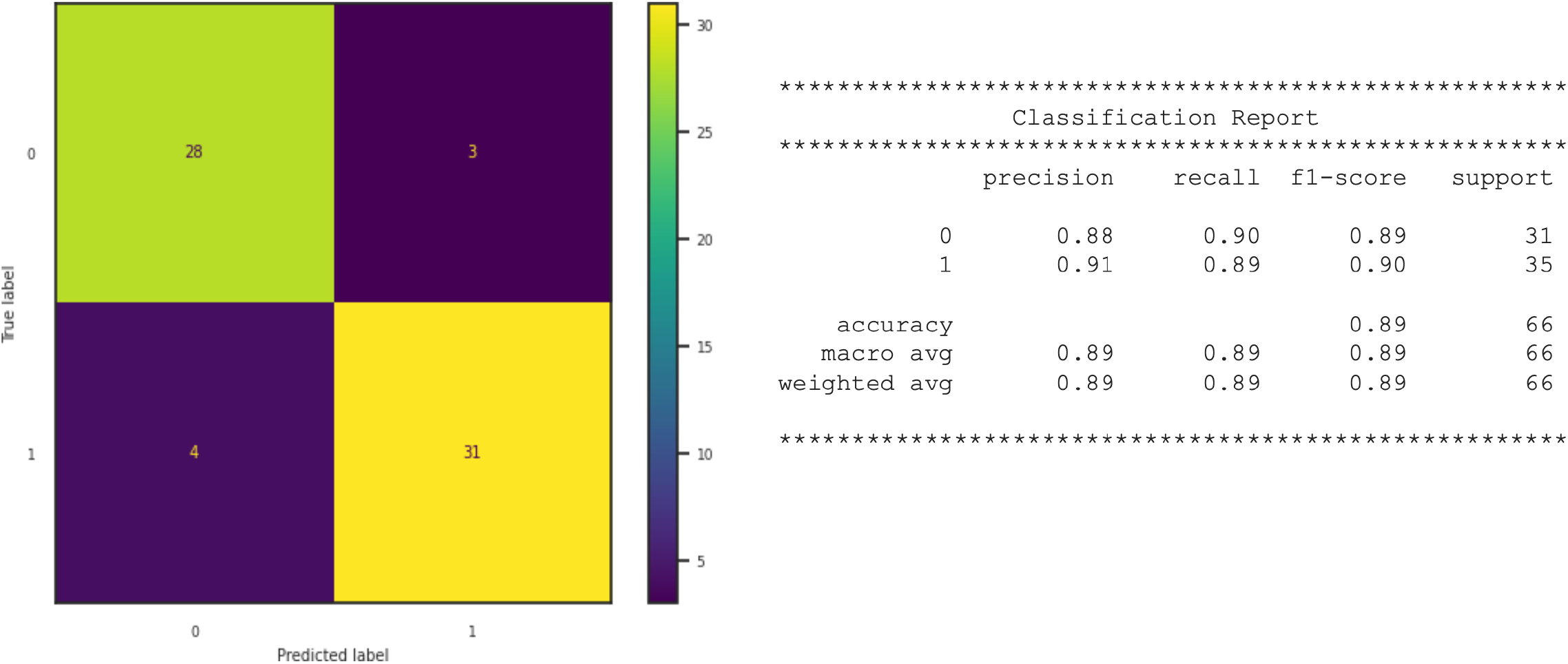
Random Forest confusion matrix indicating model quality.

**Table 1:** Performance of model classifiers following SMOTE rebalancing.

| Model | Accuracy | Balanced Accuracy | ROC AUC | F1 Score | Time Taken |
| --- | --- | --- | --- | --- | --- |
| RandomForestClassifier | 0.8939393939 | 0.8907834101 | 0.8907834101 | 0.8934222654 | 0.183555603 |
| ExtraTreesClassifier | 0.8787878788 | 0.8746543779 | 0.8746543779 | 0.8778841537 | 0.1532819271 |
| XGBClassifier | 0.8636363636 | 0.8622119816 | 0.8622119816 | 0.8634789362 | 0.06385040283 |
| NuSVC | 0.8636363636 | 0.8622119816 | 0.8622119816 | 0.8634789362 | 0.01609992981 |
| LGBMClassifier | 0.8484848485 | 0.8479262673 | 0.8479262673 | 0.8484848485 | 0.09533619881 |
| LabelPropagation | 0.8484848485 | 0.8442396313 | 0.8442396313 | 0.8473551922 | 0.01989364624 |
| LabelSpreading | 0.8484848485 | 0.8442396313 | 0.8442396313 | 0.8473551922 | 0.03365707397 |
| BaggingClassifier | 0.8333333333 | 0.8299539171 | 0.8299539171 | 0.8325207027 | 0.04491662979 |
| KNeighborsClassifier | 0.8333333333 | 0.8262672811 | 0.8262672811 | 0.8303030303 | 0.01888203621 |
| DecisionTreeClassifier | 0.803030303 | 0.8013824885 | 0.8013824885 | 0.8028029079 | 0.01473617554 |
| AdaBoostClassifier | 0.7575757576 | 0.7548387097 | 0.7548387097 | 0.7569023569 | 0.125483036 |
| ExtraTreeClassifier | 0.7424242424 | 0.7405529954 | 0.7405529954 | 0.7421268795 | 0.01395106316 |
| SVC | 0.7272727273 | 0.7299539171 | 0.7299539171 | 0.7272727273 | 0.01601076126 |
| SGDClassifier | 0.696969697 | 0.6921658986 | 0.6921658986 | 0.6947103844 | 0.01456356049 |
| RidgeClassifierCV | 0.696969697 | 0.6995391705 | 0.6995391705 | 0.696969697 | 0.01888251305 |
| LinearDiscriminantAnalysis | 0.6818181818 | 0.6852534562 | 0.6852534562 | 0.6814528755 | 0.01936411858 |
| RidgeClassifier | 0.6818181818 | 0.6852534562 | 0.6852534562 | 0.6814528755 | 0.01605606079 |
| LinearSVC | 0.6818181818 | 0.6834101382 | 0.6834101382 | 0.6820373656 | 0.04712438583 |
| CalibratedClassifierCV | 0.6666666667 | 0.669124424 | 0.669124424 | 0.6666666667 | 0.09294962883 |
| QuadraticDiscriminantAnalysis | 0.6515151515 | 0.6603686636 | 0.6603686636 | 0.6452849836 | 0.01543951035 |
| LogisticRegression | 0.6212121212 | 0.6225806452 | 0.6225806452 | 0.6214730543 | 0.0338010788 |
| BernoulliNB | 0.6212121212 | 0.6188940092 | 0.6188940092 | 0.6207748228 | 0.01476740837 |
| Perceptron | 0.5909090909 | 0.5958525346 | 0.5958525346 | 0.5889328063 | 0.0122988224 |
| GaussianNB | 0.5303030303 | 0.5405529954 | 0.5405529954 | 0.5174481659 | 0.01833105087 |
| PassiveAggressiveClassifier | 0.5151515152 | 0.5244239631 | 0.5244239631 | 0.5043068147 | 0.01719117165 |
| NearestCentroid | 0.5151515152 | 0.5188940092 | 0.5188940092 | 0.5138146168 | 0.01381254196 |
| DummyClassifier | 0.4696969697 | 0.5 | 0.5 | 0.3002186817 | 0.01414322853 |

### Feature importance

To determine the importance of each variable (feature) to the outcome (blood glucose level), we carried out random forest feature analysis. The importance of a feature is calculated based on how much the tree nodes that use that feature reduce impurity across all trees in the forest. The key findings show that uric acid and age are the most important features associated with elevated blood glucose (Figure 11), followed by systolic blood pressure and body mass index (BMI).

**Figure 11:**
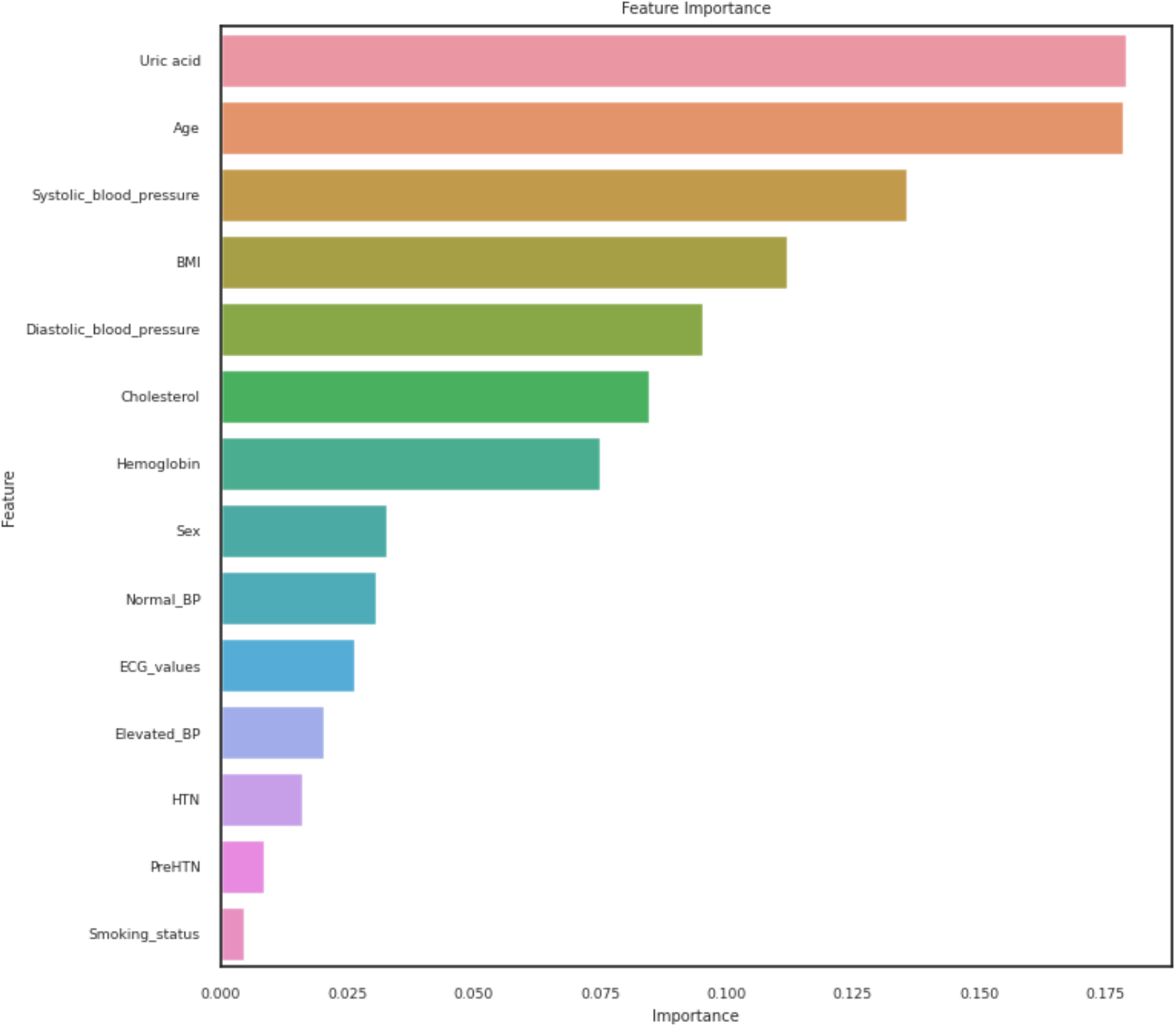
Blood glucose risk predictors.

## Discussion

Noncommunicable diseases, such as cancer, cardiovascular diseases, and diabetes, are progressively becoming the primary causes of mortality in sub-Saharan Africa [21]. This epidemiological shift is primarily attributed to limitations in implementing crucial control measures, such as prevention and early detection [1]. This research focuses on exploring key clinical indices of NCDs in asymptomatic individuals. Specifically, the study employs various machine learning algorithms to predict hyperglycemia to enable early identification of individuals at a particular risk of developing diabetes. The application of machine learning in disease prediction is now well-established for its immense potential in analyzing complex datasets and uncovering patterns that may elude human detection [22].

### Hypertension Dynamics and Age-Related Patterns

The study identified suspected hypertension in 21% of study participants, underscoring the urgency of addressing hypertension as a major health challenge in the country. Furthermore, a notable increase in the prevalence of hypertension with advancing age was observed. However, the investigation into hypertension subtypes revealed a dual phenomenon: a pronounced increase in systolic hypertension with age and a concomitant reduction in diastolic hypertension. Several factors may contribute to the observed age-related increase in systolic hypertension. Physiological changes, alterations in vascular reactivity, and lifestyle factors could play decisive roles in driving the upward trajectory of systolic blood pressure with advancing age [23, 24]. In contrast, the age-related reduction in diastolic hypertension may be associated with changes in arterial compliance, heart rate dynamics, or other physiological adaptations over the aging process [25]. Recognizing these dual dynamics holds significant clinical implications, necessitating tailored screening protocols and interventions to address the unique challenges posed by hypertension in different age groups.

Moreover, a gender disparity was observed, with systolic hypertension being more prevalent in females while diastolic hypertension was more common in males. This gender difference may be linked to heart rate variability or hormonal influences, particularly fluctuations in estrogen levels in females. Understanding how blood vessels respond to changes in pressure and the potential impact on systolic blood pressure is crucial in deciphering these gender disparities [26–28]. Therefore, tailoring screening protocols and interventions to address the unique challenges posed by hypertension in different age groups and genders is essential to mitigate the overall burden of this condition.

### Age-Dependent Patterns in ECG Findings

Electrocardiography is a pivotal tool for assessing cardiac health, and its interpretation can provide valuable insights into cardiovascular conditions. Our investigation revealed a remarkable age-dependent pattern in abnormal ECG values, reaching a peak at 70 years. Advancing age often coincides with a myriad of physiological changes, including alterations in cardiac structure and function [29–31]. A comprehensive exploration of these factors is essential for delineating the intricate relationship between aging and abnormal ECG findings.

### Prevalence of Prediabetes and Diabetes in Apparently Healthy Individuals

The global burden of diabetes is well-documented [32–34], but our investigation into supposedly healthy individuals has unearthed a concerning revelation. Despite outward appearances of health, there exists a relatively high prevalence of suspected prediabetes and diabetes in the cohort. This underscores the importance of probing beyond outward health markers to understand latent metabolic landscape [35–38]. This prompts a reevaluation of health screening protocols to incorporate metabolic parameters in apparently healthy populations. Early detection and intervention strategies should be tailored to encompass metabolic assessments, providing an opportunity for targeted preventive measures and lifestyle modifications.

### Machine Learning Algorithm Selection

In the realm of predictive modeling, selecting the most effective machine learning algorithm is paramount. Our study, aimed at evaluating various algorithms, revealed insightful findings regarding their predictive performance. Upon meticulous evaluation, Random Forest emerged as the top-performing algorithm, consistently delivering the highest accuracy among the tested models. The success of the Random Forest algorithm can be attributed to its ensemble learning nature [39, 40], which harnesses the collective power of multiple decision trees. This enables robustness against overfitting, enhanced generalization, and effective handling of complex datasets with diverse features. The observed superiority of Random Forest in our study has profound implications for future applications, suggesting its applicability across diverse datasets and underscoring its potential as a reliable choice for achieving high predictive accuracy.

### Feature Importance Analysis: Uric Acid and Age as Predictors

In the pursuit of understanding the intricate determinants of hyperglycemia, our study employed a robust feature importance analysis, with compelling results showcasing uric acid and age as the most influential predictors. Uric acid’s prominence as a predictor of hyperglycemia adds a unique dimension to our understanding of metabolic health. While traditionally associated with conditions like gout, our findings suggest a potential link between uric acid levels and hyperglycemia, urging further exploration into the underlying physiological mechanisms. The identification of age as a key predictor aligns with existing knowledge regarding the age-associated risk of hyperglycemia [40–42]. Our findings reinforce the significance of age as a robust indicator, reflecting the cumulative impact of aging processes on metabolic health and glucose regulation. The recognition of uric acid and age as pivotal predictors holds significant clinical implications. Healthcare practitioners can leverage these findings to enhance risk assessment strategies for hyperglycemia. Incorporating uric acid measurements and age considerations into routine screenings may facilitate early identification of individuals at heightened risk, enabling proactive interventions. While our study sheds light on the importance of uric acid and age, further research is warranted to unravel the intricate relationships and mechanisms underlying these associations. Longitudinal studies exploring the dynamic interplay between uric acid, age and hyperglycemia can deepen our understanding and inform targeted interventions.

### Limitations and Future Directions

While our study provides valuable insights into predicting hyperglycemia using machine learning in undiagnosed individuals, it is essential to acknowledge certain limitations that may impact interpretation. The size of our cohort may limit the generalizability of our results. A larger and more diverse sample could enhance the external validity of the predictive model. Furthermore, the study did not account for potential variations in clinical practice, including differences in diagnostic criteria. For instance, the study did not take into consideration orthostatic hypotension, a fall in SBP of at least 20 mm Hg or a DBP fall of at least 10 mm Hg within three minutes of standing, especially in older individuals [14]. Although seats were provided to participants, we could not accurately document how long participants had been standing before attending the screening. Besides, phenomena such as postprandial hypotension (a reduction in BP after meals, a common cause of syncope and falls in healthy and hypertensive elderly individuals), circadian BP variability, and white-coat (non-sustained) hypertension, especially in the elderly were not factored into the analyses [43–45]. Incorporating standardized criteria across diverse healthcare settings could enhance our model’s clinical applicability. In addition, the study did not dissect the influence of ethnicity and genetics on hyperglycemia [46, 47]. Future research could explore these aspects to provide a more comprehensive understanding of predictive factors. Since the dataset primarily comprises information from a specific geographic location or demographic group, extrapolating the findings to other populations requires caution as regional variations in lifestyle, genetics, and healthcare practices may influence the performance of the predictive model. Moreover, the cross-sectional nature of our study limits our ability to establish causation or assess changes over time. As such, longitudinal studies could be beneficial to understand the dynamic nature of hyperglycemia predictors. The model’s performance was evaluated on the same dataset used for training, raising the potential for overfitting. External validation on an independent dataset is required to assess its generalizability and reliability in real-world scenarios. Lastly, the importance of a feature in a Random Forest model does not necessarily mean a causal relationship and other models might find different results if additional features are introduced. In essence, future approaches are expected to accommodate more features and larger datasets. This will account for the deployment of built and containerized models as publicly accessible web apps. In all, this present study has expounded the potential of machine learning for early disease detection, risk assessment strategies, proactive interventions and targeted therapeutic design.

## Conclusions

This study makes a substantial contribution to the expanding domain of predictive modeling and offers promising implications for enhancing early detection and personalized risk assessment, particularly in the context of hyperglycemia and its potential association with diabetes. The research has not only brought to light the prevalence of undiagnosed hypertension, isolated systolic and diastolic hypertension but has also highlighted factors associated with elevated blood glucose within the population. The findings of this study emphasize the significance of regular screening, effective intervention strategies and targeted therapeutic designs. Collectively, the results contribute to the overarching endeavor to enhance healthcare outcomes through proactive and tailored approaches.

## Data Availability

All relevant data are within the manuscript and its Supporting File.The Google Colab Python Script used for data analysis and machine learning has been deposited in our GitHub page https://github.com/oyebolakolapo/Machine-Learning-Prediction-of-Elevated-Blood-Glucose-in-a-Cohort-of-Apparently-Healthy-Adults

## Acknowledgements

The authors appreciate the study participants and Ijede Community Leaders for their cooperation during the screening exercise.

## Contributions

KO conceived and designed the study. KO, FL, AO, BE, YA and OA implemented field study. KO, FL and BE carried out laboratory experiments. KO carried out data analysis, including machine learning. KO drafted the manuscript. KO, OA and BS edited the manuscript. All authors read and approved the final manuscript.

## Funding

Kolapo Oyebola was supported by APTI-18-07 Grant by the African Academy of Sciences in partnership with Bill and Melinda Gates Foundation; and a Fogarty Emerging Global Leader Grant (NIH-K43TW011926) from the US National Institutes of Health. The funders had no role in study design, data collection and analysis, decision to publish or preparation of the manuscript.

## Consent for publication

Not applicable.

## Competing interests

The authors declare no competing interests.

## Notes

### Competing Interest Statement

The authors have declared no competing interest.

### Author Declarations

Ethical approval was obtained from the Institutional Review Board of the Nigerian Institute of Medical Research (IRB/21/074)

## References

1. Bigna, J.J. and J.J. Noubiap, The rising burden of non-communicable diseases in sub-Saharan Africa. Lancet Glob Health, 2019. 7(10): p. e1295–e1296.

2. Cross, S.H., et al., Rural-Urban Differences in Cardiovascular Mortality in the US, 1999-2017. JAMA, 2020. **323**(18): p. 1852-1854.

3. Turecamo, S.E., et al., Association of Rurality With Risk of Heart Failure. JAMA Cardiology, 2023. 8(3): p. 231–239.

4. Khayat, S., et al., Lifestyles in suburban populations: A systematic review. Electron Physician, 2017. 9(7): p. 4791–4800.

5. Kolié, D., et al., Increasing the availability of health workers in rural sub-Saharan Africa: a scoping review of rural pipeline programmes. Human Resources for Health, 2023. 21(1): p. 20.

6. Ngene, N.C., O.P. Khaliq, and J. Moodley, Inequality in health care services in urban and rural settings in South Africa. Afr J Reprod Health, 2023. 27(5s): p. 87–95.

7. Jane Ling, M.Y., N. Ahmad, and A.N. Aizuddin, Risk perception of non-communicable diseases: A systematic review on its assessment and associated factors. PLoS One, 2023. 18(6): p. e0286518.

8. Tohidinezhad, F., et al., The burden and predisposing factors of non-communicable diseases in Mashhad University of Medical Sciences personnel: a prospective 15-year organizational cohort study protocol and baseline assessment. BMC Public Health, 2020. 20(1): p. 1637.

9. Alanazi, R., Identification and Prediction of Chronic Diseases Using Machine Learning Approach. J Healthc Eng, 2022. 2022: p. 2826127.

10. Park, D.J., et al., Development of machine learning model for diagnostic disease prediction based on laboratory tests. Scientific Reports, 2021. 11(1): p. 7567.

11. Uddin, S., et al., Comparing different supervised machine learning algorithms for disease prediction. BMC Medical Informatics and Decision Making, 2019. 19(1): p. 281.

12. Keohane, E.M., L. Smith, and J.M. Walenga, Rodak’s Hematology - E-Book: Rodak’s Hematology - E-Book. 2015: Elsevier Health Sciences.

13. Yousefi, M., et al., Association of consumption of excess hard water, body mass index and waist circumference with risk of hypertension in individuals living in hard and soft water areas. Environ Geochem Health, 2019. 41(3): p. 1213–1221.

14. Tan, J.L. and K. Thakur, *Systolic Hypertension*, in *StatPearls*. 2023: Treasure Island (FL).

15. Whelton, P.K., et al., 2017 ACC/AHA/AAPA/ABC/ACPM/AGS/APhA/ASH/ASPC/NMA/PCNA Guideline for the Prevention, Detection, Evaluation, and Management of High Blood Pressure in Adults: Executive Summary: A Report of the American College of Cardiology/American Heart Association Task Force on Clinical Practice Guidelines. Circulation, 2018. 138(17): p. e426-e483.

16. Diagnosis and classification of diabetes mellitus. Diabetes Care, 2010. 33 Suppl 1(Suppl 1): p. S62-9.

17. Pedregosa, F., et al., Scikit-learn: Machine Learning in Python. J. Mach. Learn. Res., 2011. 12(null): p. 2825–2830.

18. N. V. Chawla, K.W.B., L. O. Hall, W. P. Kegelmeyer, SMOTE: Synthetic Minority Over-sampling Technique. Journal of Artifical Intelligence Research, 2002. 16: p. 321–357.

19. Buyya, R., et al., Computational Intelligence and Data Analytics: Proceedings of ICCIDA 2022. 2022: Springer Nature Singapore.

20. Lathkar, M., High-Performance Web Apps with FastAPI. Apress Berkeley, CA, 2023.

21. Katende, D., et al., Medium-to-long term sustainability of a health systems intervention to improve service readiness and quality of non-communicable disease (NCD) patient care and experience at primary care settings in Uganda. BMC Health Serv Res, 2023. 23(1): p. 1022.

22. Davenport, T. and R. Kalakota, The potential for artificial intelligence in healthcare. Future Healthc J, 2019. 6(2): p. 94–98.

23. Sharifi-Rad, J., et al., *Diet,* Lifestyle and Cardiovascular Diseases: Linking Pathophysiology to Cardioprotective Effects of Natural Bioactive Compounds. Int J Environ Res Public Health, 2020. 17(7).

24. Liu, R., et al., Systolic Blood Pressure Trajectories and the Progression of Arterial Stiffness in Chinese Adults. Int J Environ Res Public Health, 2022. 19(16).

25. Singh, J.N., et al., Physiology, Blood Pressure Age Related Changes, in *StatPearls*. 2023, StatPearls Publishing Copyright © 2023, StatPearls Publishing LLC.: Treasure Island (FL).

26. Song, J.J., et al., Gender Differences in Hypertension. J Cardiovasc Transl Res, 2020. 13(1): p. 47–54.

27. Wu, J., B. Jiao, and Y. Fan, Urbanization and systolic/diastolic blood pressure from a gender perspective: Separating longitudinal from cross-sectional association. Health Place, 2022. 75: p. 102778.

28. Midtbø, H. and E. Gerdts, Sex disparities in blood pressure development: time for action. Eur J Prev Cardiol, 2022. 29(1): p. 178–179.

29. Fleg, J.L. and D.E. Forman, Aging Changes in Cardiovascular Structure and Function, in Handbook of Cardiovascular Behavioral Medicine, S.R. Waldstein, et al., Editors. 2022, Springer New York: New York, NY. p. 127-162.

30. Fleg, J.L. and J. Strait, Age-associated changes in cardiovascular structure and function: a fertile milieu for future disease. Heart Fail Rev, 2012. 17(4-5): p. 545–54.

31. Hacker, T.A., et al., Age-related changes in cardiac structure and function in Fischer 344 × Brown Norway hybrid rats. American Journal of Physiology-Heart and Circulatory Physiology, 2006. 290(1): p. H304–H311.

32. King, H., R.E. Aubert, and W.H. Herman, Global burden of diabetes, 1995-2025: prevalence, numerical estimates, and projections. Diabetes Care, 1998. 21(9): p. 1414-31.

33. Herman, W.H., The Global Burden of Diabetes: An Overview, in *Diabetes Mellitus in Developing Countries and Underserved Communities, S. Dagogo-Jack, Editor*. 2017, Springer International Publishing: Cham. p. 1-5.

34. Global, regional, and national burden of diabetes from 1990 to 2021, with projections of prevalence to 2050: a systematic analysis for the Global Burden of Disease Study 2021. Lancet, 2023. 402(10397): p. 203-234.

35. Huang, P.L., A comprehensive definition for metabolic syndrome. Dis Model Mech, 2009. 2(5-6): p. 231–7.

36. Rafaqat, S., et al., Biomarkers of Metabolic Syndrome: Role in Pathogenesis and Pathophysiology Of Atrial Fibrillation. J Atr Fibrillation, 2021. 14(2): p. 20200495.

37. Srikanthan, K., et al., Systematic Review of Metabolic Syndrome Biomarkers: A Panel for Early Detection, Management, and Risk Stratification in the West Virginian Population. Int J Med Sci, 2016. 13(1): p. 25–38.

38. Madhusoodanan, J., Searching for Better Biomarkers for Metabolic Syndrome. ACS Central Science, 2022. 8(6): p. 682–685.

39. Schonlau, M. and R.Y. Zou, The random forest algorithm for statistical learning. The Stata Journal, 2020. 20(1): p. 3–29.

40. Ghaffar Nia, N., E. Kaplanoglu, and A. Nasab, Evaluation of artificial intelligence techniques in disease diagnosis and prediction. 2023. 3(1).

41. Longo, M., et al., Diabetes and Aging: From Treatment Goals to Pharmacologic Therapy. Frontiers in Endocrinology, 2019. 10.

42. Yan, Z., et al., The Interaction Between Age and Risk Factors for Diabetes and Prediabetes: A Community-Based Cross-Sectional Study. Diabetes Metab Syndr Obes, 2023. 16: p. 85–93.

43. Nuredini, G., et al., Current status of white coat hypertension: where are we? Ther Adv Cardiovasc Dis, 2020. 14: p. 1753944720931637.

44. Franklin, S.S., et al., White-Coat Hypertension. Hypertension, 2013. 62(6): p. 982–987.

45. Luciano, G.L., M.J. Brennan, and M.B. Rothberg, Postprandial hypotension. Am J Med, 2010. 123(3): p. 281.e1–6.

46. Ali, O., Genetics of type 2 diabetes. World J Diabetes, 2013. 4(4): p. 114–23.

47. Li, C., et al., Glucose metabolism-related gene polymorphisms as the risk predictors of type 2 diabetes. Diabetology & Metabolic Syndrome, 2020. 12(1): p. 97.

