## Supplemental File for "Machine Learning-Based Hyperglycemia Prediction: Enhancing Risk Assessment in a Cohort of Undiagnosed Individuals"

#### Supplementary Methods

##### Train-test dataset splitting

This code is used to split the data into training and test sets for a machine learning model (https://github.com/oyebolakolapo/Machine-Learning-Prediction-of-Elevated-Blood-Glucose-in-a-Cohort-of-Apparently-Healthy-Adults). First, the code separates the features (x) and target (y) variables from the original dataset bank_data using the drop() method. The drop() method is used to remove a specified column from the dataset. In this case, the column 'y' is removed from the dataset and assigned to the variable x. The column 'y' is assigned to the variable y. Next, the train_test_split() function from the scikit-learn library is used to split the data into training and test sets. The function takes four arguments: the scaled features (X), the target (y), the test size (0.2 in this case), and a random state (which is not specified in this code). The test size argument specifies the proportion of the data that should be used for testing the model. In this case, 20% of the data is used for testing and 80% is used for training. The function returns four variables: X_train, X_test, y_train, and y_test. These variables contain the training and test sets for the features and target variables. The training sets are used to train the machine learning model, and the test sets are used to evaluate the performance of the model.

**Data cleaning steps and outcomes**

The original dataset contained 22 columns and 195 rows, eight continuous variable and 14 categorical variables. The dataset was checked for null values with seaborn heatmap used as a visual scan. For continuous variables, mean values were imputed into missing cells while modes were adopted for categorical variables. Subsequently, "Normal_glucose" and Normal_ECG_Values" were recoded and 'High" and 'Normal" values were replaced with "0" and "1". Columns "Normal_glucose" was then renamed as "Blood_glucose", while "Normal_ECG_Values" was renamed as "ECG_values". No duplicate rows were observed. We checked and found no outliers. Next, we had a cursory view of the target variable (Blood_glucose) to check the distribution of the two outcomes "high" and "normal" recoded as "1" and "0" respectively. Count of "0" was 83.59% and 16.4% for "1". Data is not balanced hence we applied SMOTE technique much later before deploying machine learning models. Meanwhile, we interrogated how the features compared in a correlation matrix.

### Supplementary Figures

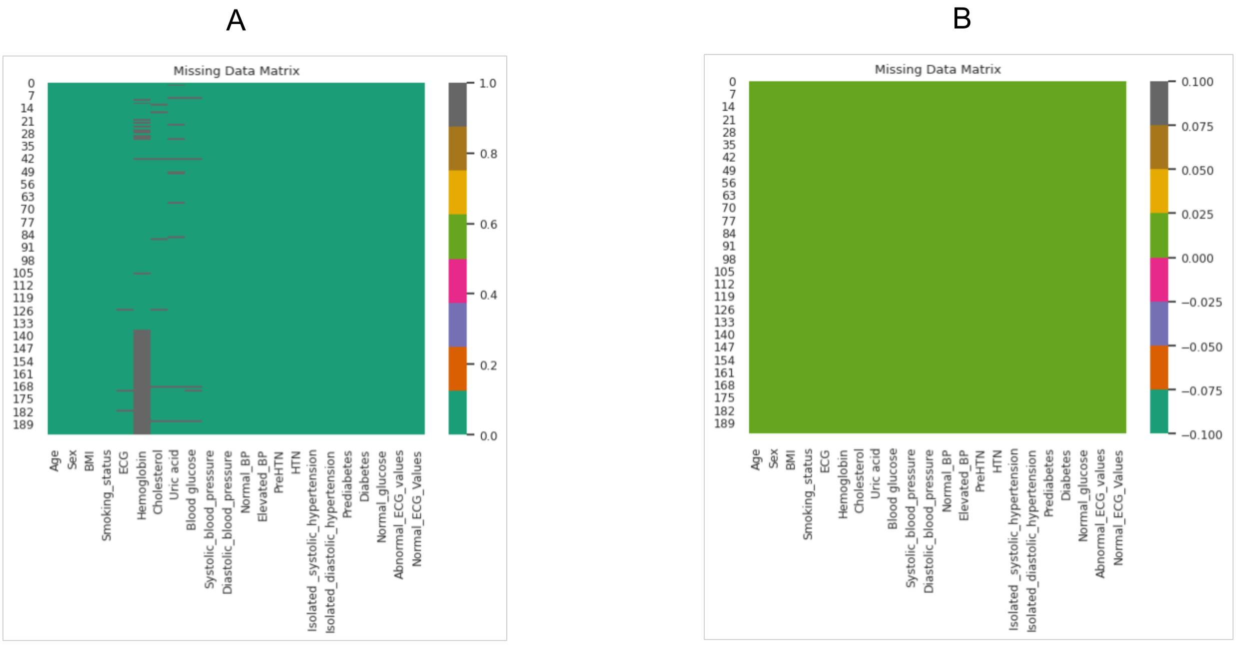


Supplementary Figure 1: Dataset overview before (A) and after (B) missing values were treated.

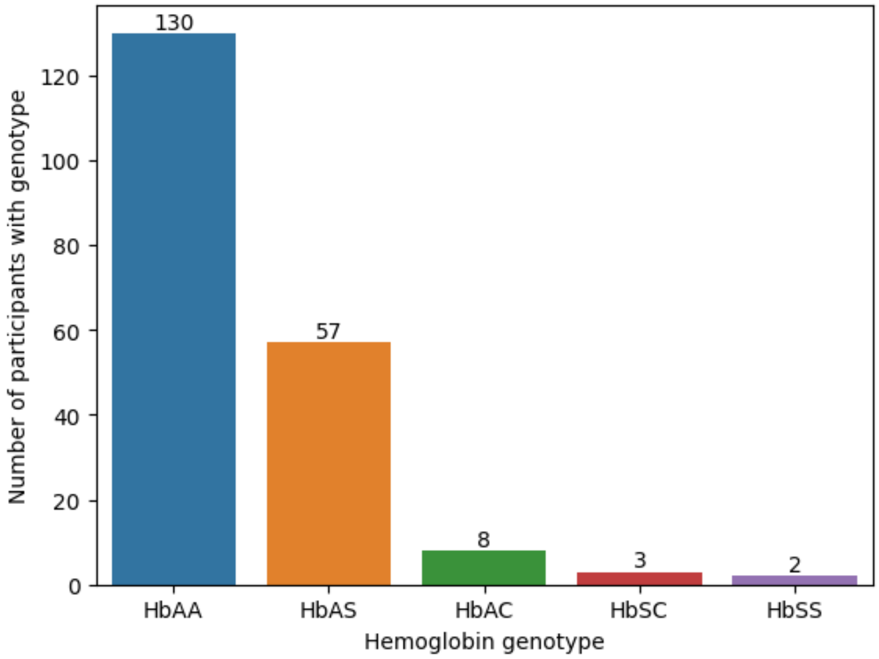


Supplementary Figure 2: Hemoglobin variants in the cohort

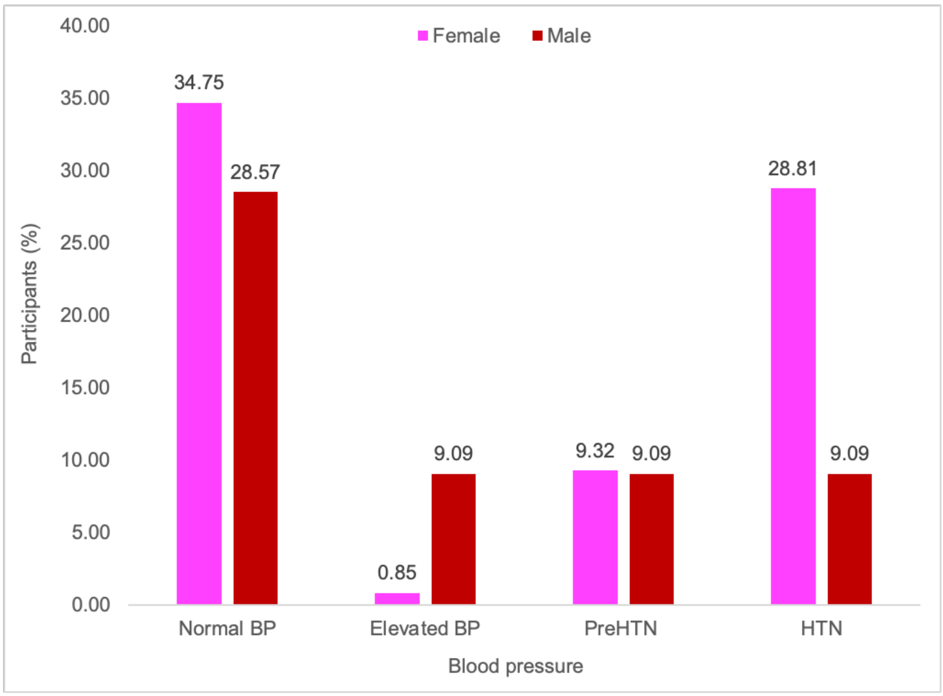

Supplementary Figure 3: Blood pressure values recorded in the cohort

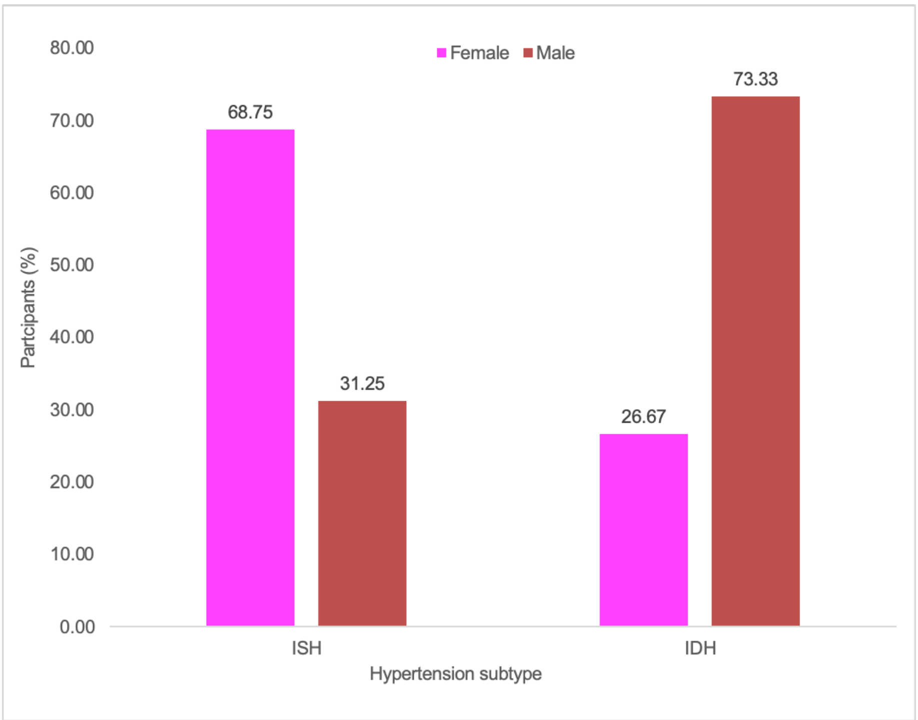


Supplementary Figure 4: Prevalence of isolated systolic hypertension (ISH) and isolated diastolic hypertension (IDH) in the cohort

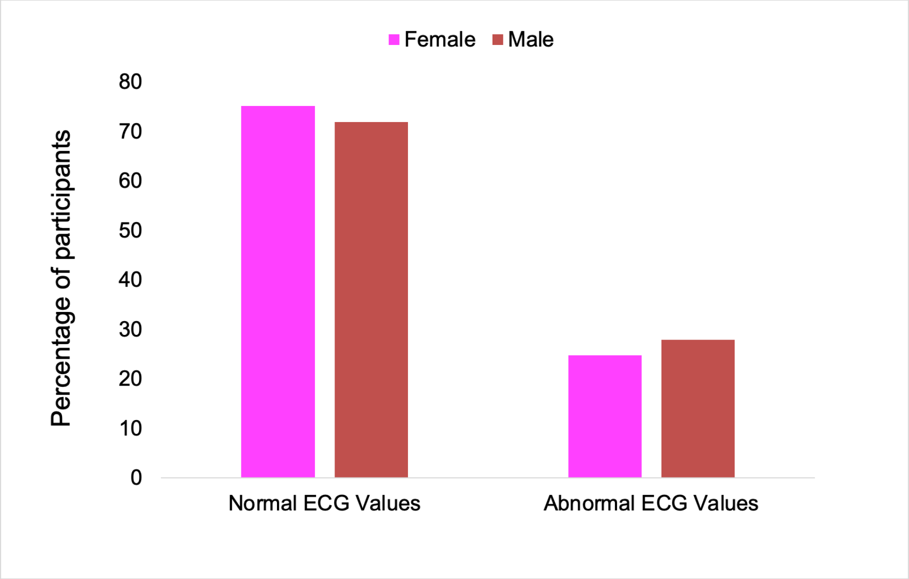


Supplementary Figure 5: Gender-based electrocardiogram (ECG) analysis

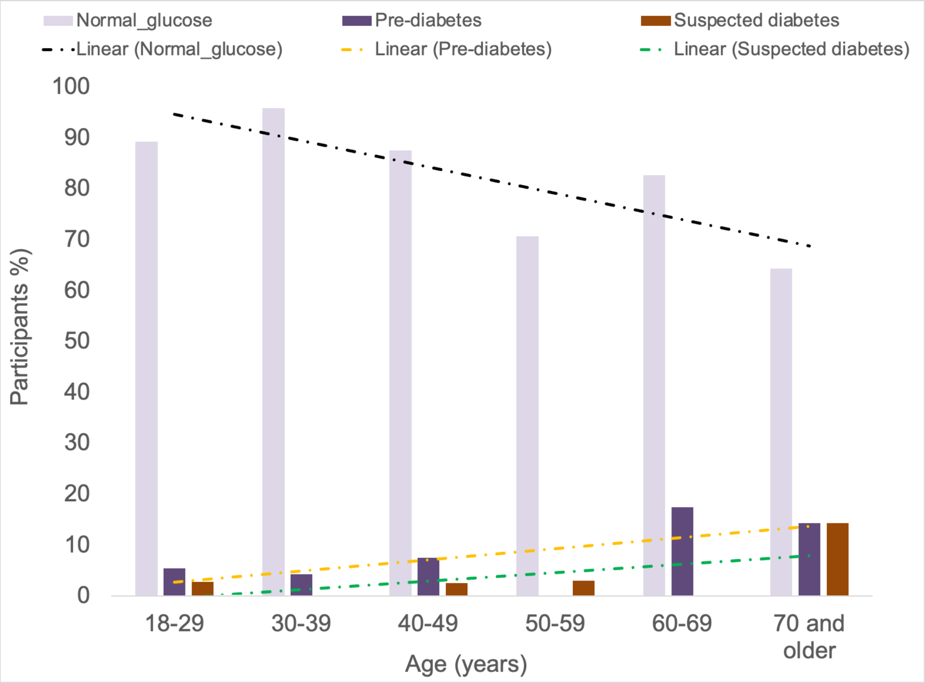


Supplementary Figure 6: Frequency of high blood glucose with age

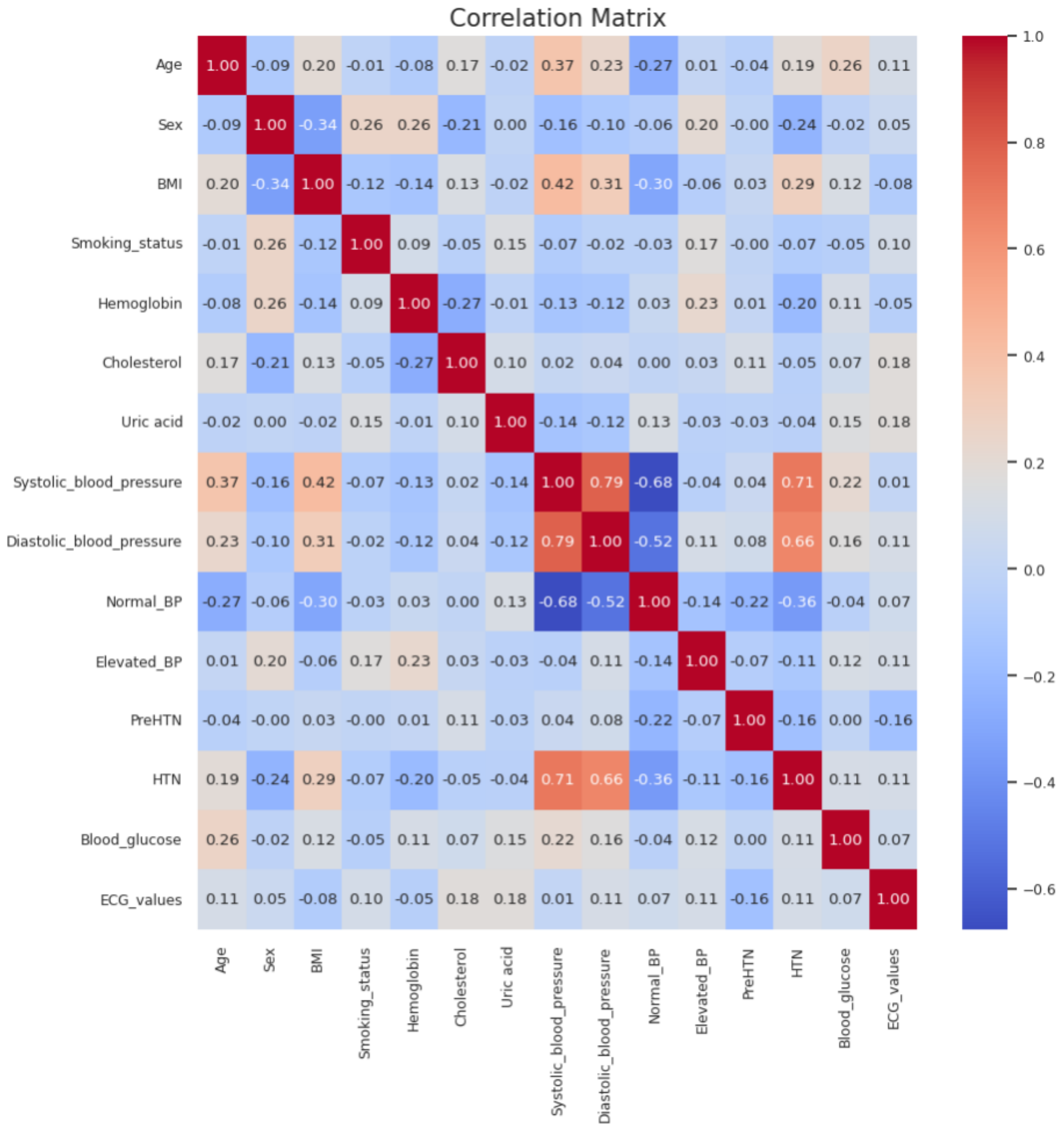


Supplementary Figure 7: Correlation matrix of variables contained in the dataset.

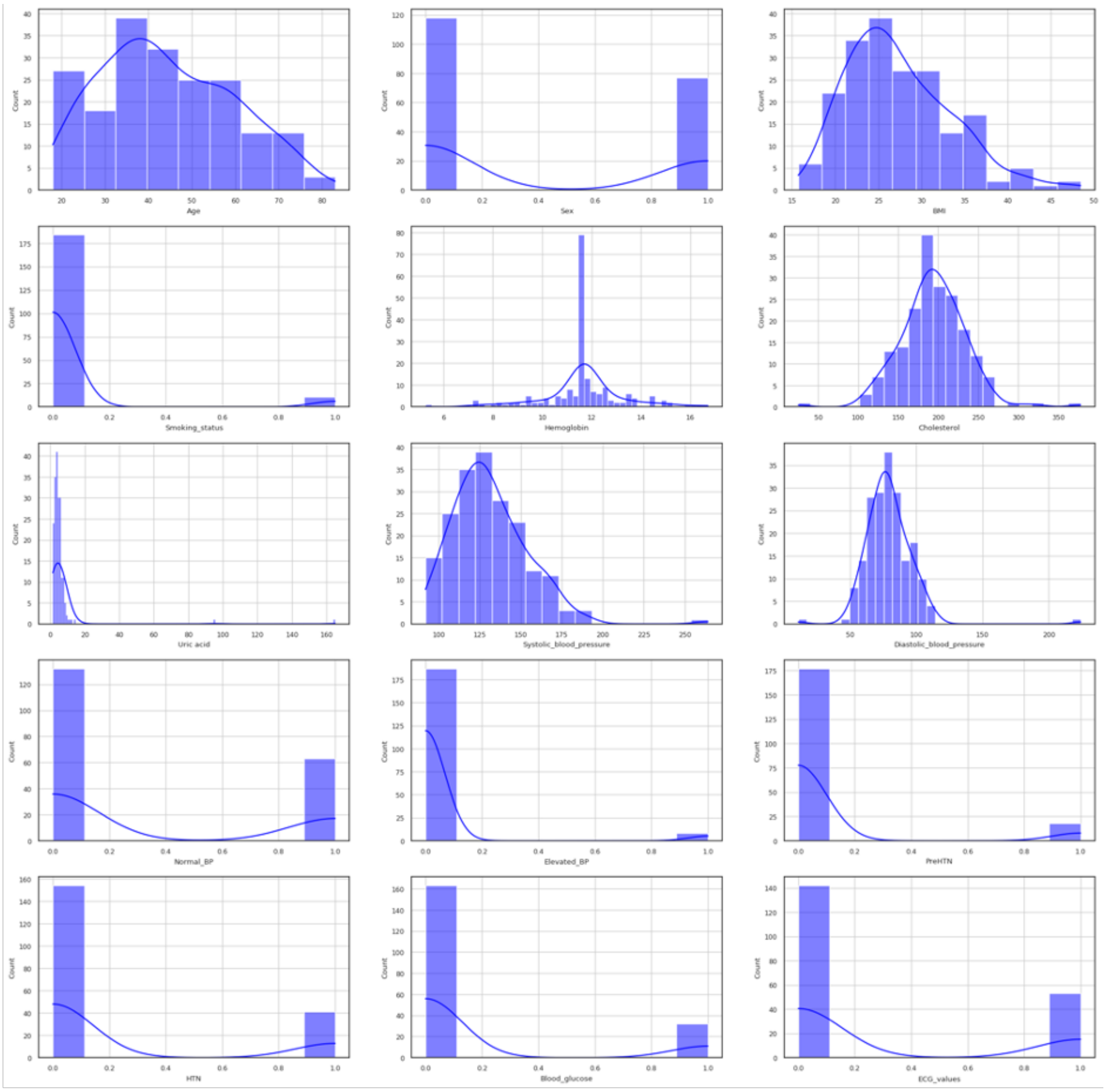


Supplementary Figure 8: Distribution of variables in cleaned dataset

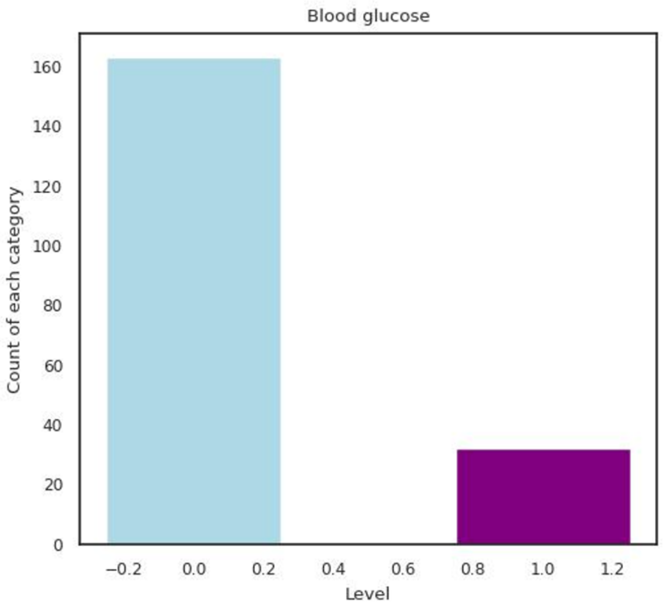


Supplementary Figure 9: Class imbalance in the outcome (blood glucose level) variable. 83.6% of the participants had normal blood glucose {0} while 16.4% had high blood glucose level {1}

**Supplementary Table 1: Model performance report before SMOTE rebalancing**

| Model | Accuracy | Balanced Accuracy | ROC AUC | F1 Score | Time Taken |
|---|---|---|---|---|---|
| RandomForestClassifier | 0.8205128205 | 0.5 | 0.5 | 0.7396171903 | 0.6792166233 |
| DummyClassifier | 0.8205128205 | 0.5 | 0.5 | 0.7396171903 | 0.1113314629 |
| AdaBoostClassifier | 0.8205128205 | 0.5558035714 | 0.5558035714 | 0.7771584293 | 0.4256854057 |
| SVC | 0.8205128205 | 0.5 | 0.5 | 0.7396171903 | 0.05286502838 |
| BernoulliNB | 0.8205128205 | 0.6116071429 | 0.6116071429 | 0.8000556657 | 0.04009056091 |
| CalibratedClassifierCV | 0.7948717949 | 0.484375 | 0.484375 | 0.7267399267 | 0.2218351364 |
| KNeighborsClassifier | 0.7948717949 | 0.484375 | 0.484375 | 0.7267399267 | 0.06800198555 |
| BaggingClassifier | 0.7948717949 | 0.484375 | 0.484375 | 0.7267399267 | 0.2022051811 |
| PassiveAggressiveClassifier | 0.7948717949 | 0.6517857143 | 0.6517857143 | 0.7948717949 | 0.0400686264 |
| QuadraticDiscriminantAnalysis | 0.7692307692 | 0.5245535714 | 0.5245535714 | 0.7429287131 | 0.05598902702 |
| ExtraTreesClassifier | 0.7692307692 | 0.46875 | 0.46875 | 0.7134894091 | 1.009959698 |
| RidgeClassifier | 0.7692307692 | 0.46875 | 0.46875 | 0.7134894091 | 0.02961349487 |
| RidgeClassifierCV | 0.7692307692 | 0.46875 | 0.46875 | 0.7134894091 | 0.03412246704 |
| XGBClassifier | 0.7692307692 | 0.46875 | 0.46875 | 0.7134894091 | 0.3984851837 |
| LGBMClassifier | 0.7692307692 | 0.46875 | 0.46875 | 0.7134894091 | 0.1343479156 |
| LinearDiscriminantAnalysis | 0.7435897436 | 0.5089285714 | 0.5089285714 | 0.7261072261 | 0.1002953053 |
| GaussianNB | 0.7435897436 | 0.5089285714 | 0.5089285714 | 0.7261072261 | 0.06342744827 |
| SGDClassifier | 0.7435897436 | 0.5647321429 | 0.5647321429 | 0.7435897436 | 0.05346345901 |
| LinearSVC | 0.7435897436 | 0.5089285714 | 0.5089285714 | 0.7261072261 | 0.07059240341 |
| Perceptron | 0.7179487179 | 0.5491071429 | 0.5491071429 | 0.7251119251 | 0.08241343498 |
| ExtraTreeClassifier | 0.7179487179 | 0.5491071429 | 0.5491071429 | 0.7251119251 | 0.09578275681 |
| LogisticRegression | 0.7179487179 | 0.4375 | 0.4375 | 0.6858017604 | 0.07524013519 |
| DecisionTreeClassifier | 0.6923076923 | 0.5334821429 | 0.5334821429 | 0.7065756824 | 0.05602097511 |
| LabelSpreading | 0.6153846154 | 0.4308035714 | 0.4308035714 | 0.6398635116 | 0.09875822067 |
| NearestCentroid | 0.6153846154 | 0.5982142857 | 0.5982142857 | 0.65916692 | 0.08929729462 |
| LabelPropagation | 0.5897435897 | 0.4151785714 | 0.4151785714 | 0.6216524217 | 0.06722593307 |

**Hyperparameter optimization before SMOTE rebalancing**

| | precision | recall | f1-score | support |
|---|---|---|---|---|
| 0 | 0.82 | 1.00 | 0.90 | 32 |
| 1 | 0.00 | 0.00 | 0.00 | 7 |
| accuracy | | | 0.82 | 39 |
| macro avg | 0.41 | 0.50 | 0.45 | 39 |
| weighted avg | 0.67 | 0.82 | 0.74 | 39 |

Best Params: {'max_depth': 30, 'min_samples_leaf': 1, 'min_samples_split': 2}
Best F1 Score: 0.15
Test F1 Score: 0.8205128205128205

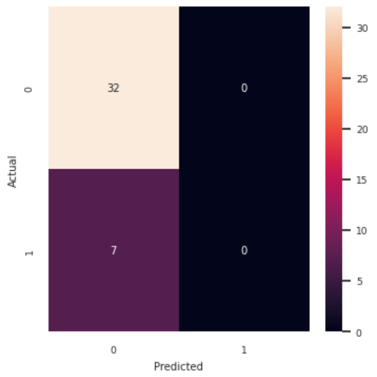


**Hyperparameter optimization after SMOTE rebalancing**

| | precision | recall | f1-score | support |
|---|---|---|---|---|
| 0 | 0.87 | 0.87 | 0.87 | 31 |
| 1 | 0.89 | 0.89 | 0.89 | 35 |
| accuracy | | | 0.88 | 66 |
| macro avg | 0.88 | 0.88 | 0.88 | 66 |
| weighted avg | 0.88 | 0.88 | 0.88 | 66 |

Best Params: {'max_depth': 10, 'min_samples_leaf': 1, 'min_samples_split': 2}
Best F1 Score: 0.8730174729485075
Test F1 Score: 0.8787878787878788

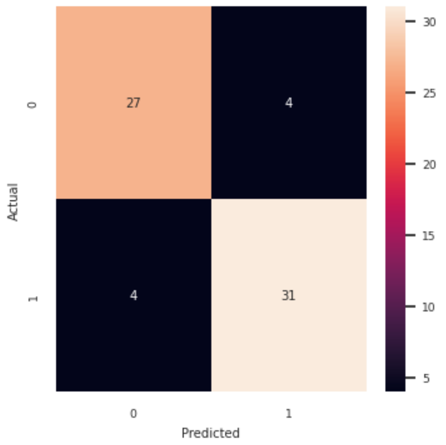
